# Differential features of the fifth wave of COVID-19 associated with vaccination and the Delta variant in a reference hospital in Catalonia, Spain

**DOI:** 10.1101/2021.10.14.21264933

**Authors:** Simona Iftimie, Ana F. López-Azcona, Maria José Lozano-Olmo, Anna Hernández-Aguilera, Salvador Sarrà-Moretó, Jorge Joven, Jordi Camps, Antoni Castro

## Abstract

Since the beginning of the COVID-19 pandemic, Spain has suffered five waves of infection, the latter being related to the expansion of the Delta variant and with a high incidence. A vaccination campaign began in December 2020 and by the end of the fifth wave 77.3% of people had been fully vaccinated. Understanding the impact of these new characteristics on COVID-19 is essential for public health strategies. Our objective was to ascertain any differences in the characteristics and outcomes of hospitalized patients during that period compared to previous waves. We found that patients in the fifth wave were considerably younger than before and the mortality rate fell from 22.5 to 2.0%. Admissions to the Intensive Care Unit decreased from 10 to 2%. Patients in the fifth wave had fewer comorbidities, and the age of the patients who died was higher than those who survived. Our results show a marked improvement in patient outcomes in the fifth wave, suggesting success of the vaccination campaign despite the explosion in cases due to the Delta variant.

## Introduction

Coronavirus disease-19 (COVID-19), produced by infection with severe acute respiratory syndrome coronavirus 2 (SARS-CoV-2) has so far affected 229 million people worldwide, causing 4.6 million deaths, according to figures reported by the WHO in September 2021^1^. Since the beginning of the pandemic, Spain has suffered five periods, or waves, of infection. The first wave of the pandemic began in March 2020 and lasted through to June 21, 2020, during which a strict curfew and mandatory home isolation of the entire population was implemented, with the exception only of essential activities. The second wave then ran from June 22 to December 6, 2020, which coincided with a partial relaxation of restrictive measures. The third wave then spanned from December 7, 2020 to March 14, 2021, which was linked to Christmas and New Year holidays and the subsequent gatherings of families and friends. The fourth wave, from March 15 to June 19, 2021, was more like a “rebound” of the third, less intense, and with relatively few seriously ill patients, probably due to the fact that a significant percentage of the older population had by that time been vaccinated. Finally, the fifth wave ran from June 20, 2021 to September 5, 2021, linked to the appearance of the Delta variant, which had a very high incidence^2^. The Delta variant is of particular concern as its transmission and viral load are considerably higher than those of the other variants detected so far, and is associated with a lower neutralizing capacity of antibodies in vaccinated or convalescent people^3-9^. In the autonomous region of Catalonia, Spain, the Alpha variant (B.1.1.7) has been the predominant one during most of the pandemic, although a significant number of cases of Beta (B.1.351) and Gamma (P.1) variants were also reported. However, as of April 16, 2021, the incidence of the Delta variant (B.1.617.2) has increased exponentially, displacing the other variants and being responsible for around 90% of infections by mid-June^10^.

The vaccination campaign in Spain began on December 27, 2020. By the start of the fifth wave, when the Delta variant began to predominate, the number of people fully vaccinated was around 30% of the population, and at the time of writing this article, in October 2021, it is 77.3 %. The vaccines administered in our country are Comirnaty (Pfizer/BioNTech), Vaxzevria (Astra-Zeneca), Spikevax (Moderna) and Ad26.COV2-S (Janssen). The emergence of the Delta variant and the success of the vaccination campaign are recent features of the pandemic that are pulling in opposite directions, so the final outcome is hard to predict. Understanding the impact of these new characteristics on the COVID-19 burden is essential if we are to fine-tune the vaccination strategy on a global scale and be prepared for the appearance of new potentially aggressive variants.

The objective of the present study was to ascertain if there were differences in the characteristics and outcomes of the fifth wave patients admitted to our center compared to those of the previous waves.

## Methods

### Study design and participants

We conducted a prospective study on patients who had attended the *Hospital Universitari de Sant Joan* during the first, second, third and fifth waves of the pandemic, between March 14, 2020 and September 15, 2021. Ours is a reference hospital for a population of over 200,000 inhabitants, including primary care centers and residences for the elderly in the area. The criteria for patients included in this study were: those treated at our center, either hospitalized or attended to in the Emergency Department, and having had an analytical diagnosis of SARS-CoV-2 infection confirmed by RT-PCR. Tests were carried out using the VIASURE *SARS-CoV-2* Real Time PCR Detection Kit (CerTest Biotec, Zaragoza, Spain), or using the Procleix® method in a Panther automated extractor and amplifier (Grifols Laboratories, Barcelona, Spain). This study was approved by the *Comitè d’Ètica i Investigació en Medicaments* (Institutional Review Board) of *Hospital Universitari de Sant Joan* (Resolution CEIM 040/2018, amended on 16 April 2020).

### Calculation of sample size

Accepting an alpha risk of 0.05 and a beta risk of less than 0.2 in a bilateral contrast, 195 subjects in each group were needed to register a difference greater than 20 cases in the variable “number of deaths”. A follow-up loss rate of 0% had been estimated. The ARCSINUS approach was used.

### Statistical analyses

Data is given as numbers and percentages or means and standard errors. Statistical comparisons between two groups were made using the χ2 test (categorical variables) or Student’s *t* test (quantitative variables). Statistical significance was set at P ≤ 0.05. All calculations were made using the SPSS 25.0 statistical package (SPSS Inc., Chicago, IL, USA).

## Results and Discussion

The number of patients admitted to our center since the onset of the pandemic is 2,671 (1,557 survivors) and Fig. 1 shows the evolution of admissions. The first 200 patients from each wave were selected prospectively for inclusion in this study. We did not take into account the fourth wave because the number of hospitalized patients was too low (n = 135) to obtain reliable statistical results. The raw data is given as Supplementary Information (S1_File) and summarized in Table 1.

**Table 1.**
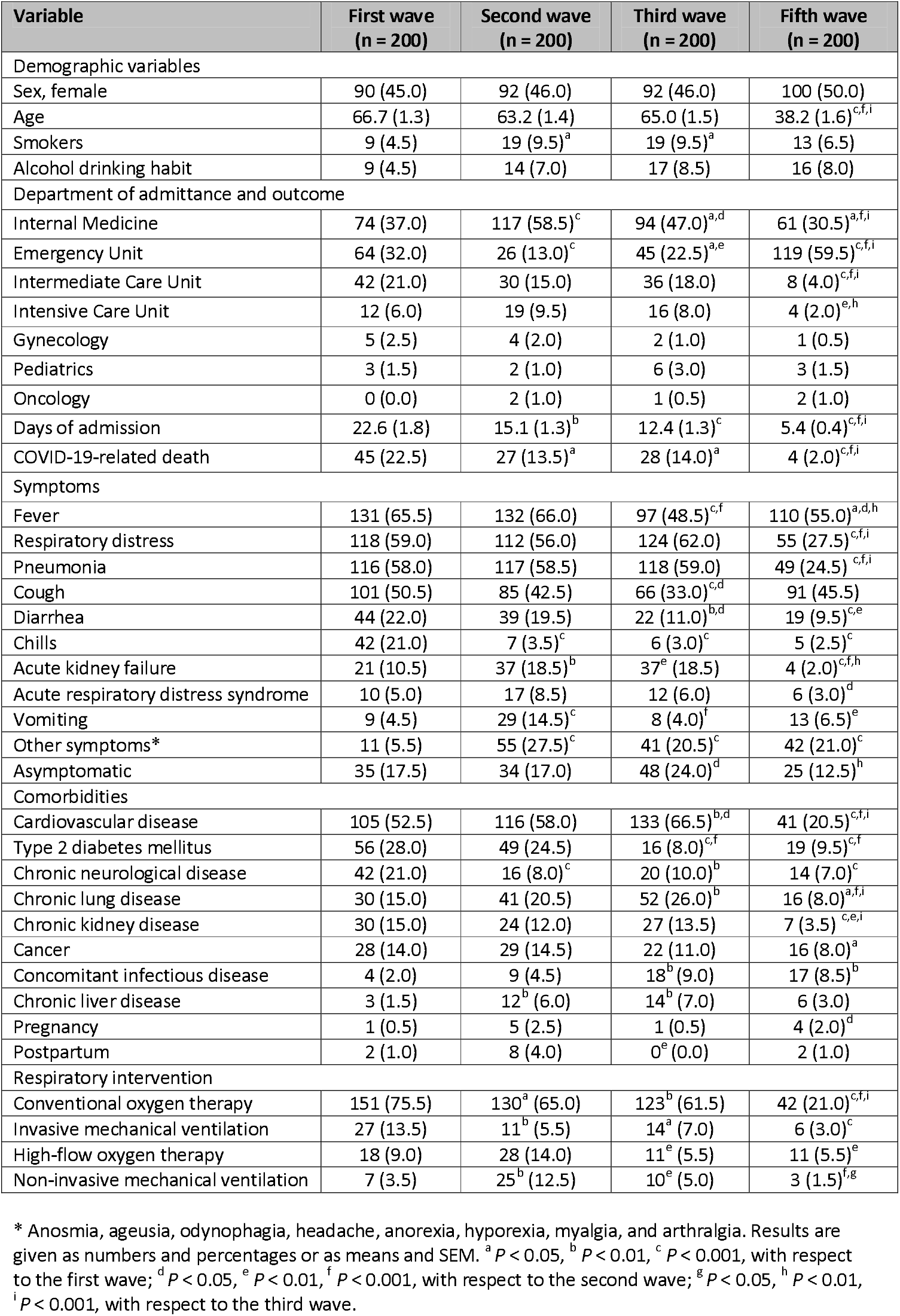
Demographic and clinical characteristics of COVID-19 patients

**Figure 1.**
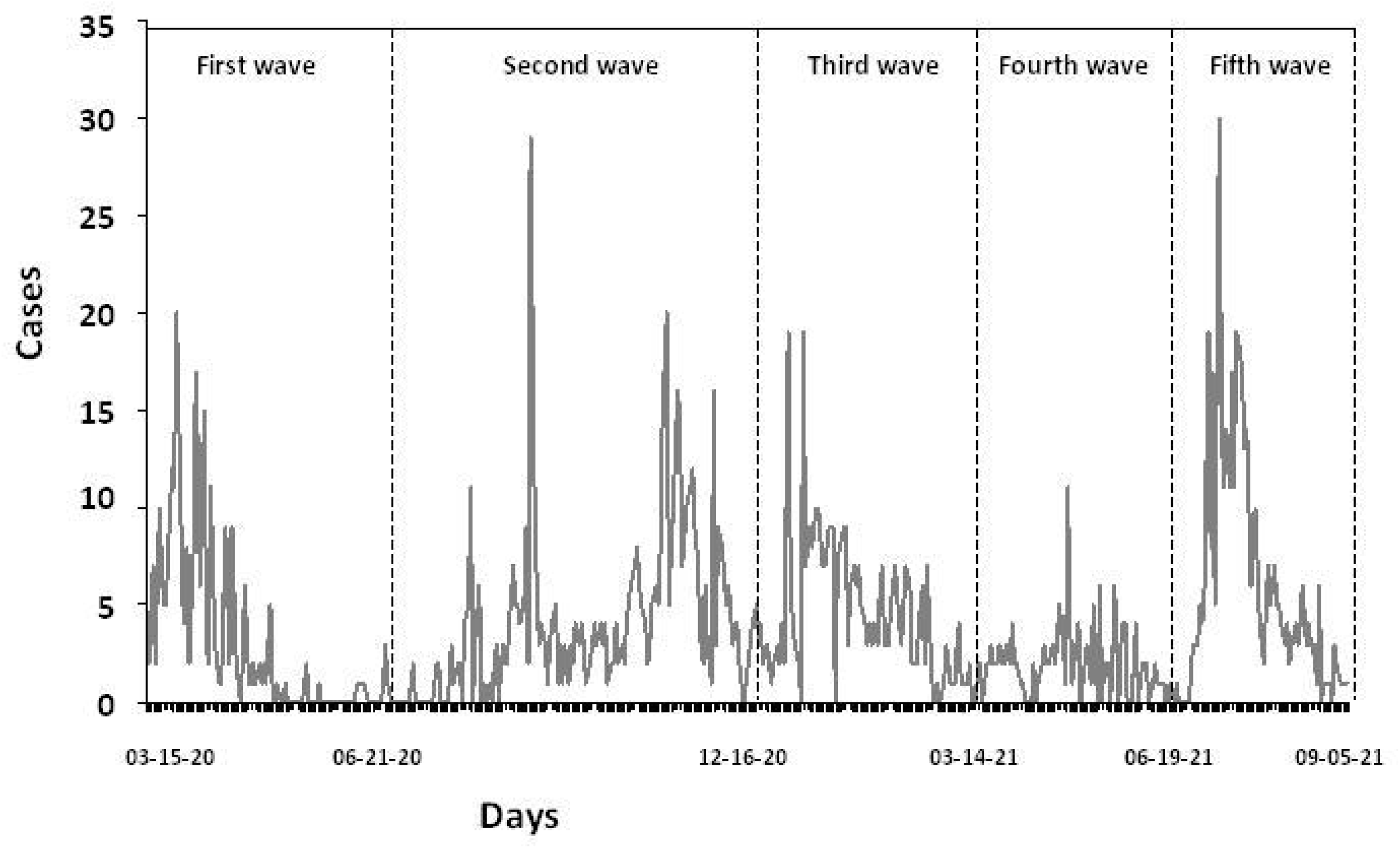
Number of patients admitted to the hospital per day from the start of the pandemic to the end of the fifth wave.

Our results for patients taken from waves 1 to 3 are similar to those reported previously by many studies around the world^11-15^. Patients had an advanced age (more than 60 years-old on average) together with age-related comorbidities, such as type 2 diabetes mellitus, hypertension and cardiovascular diseases. The proportion of deaths in the first wave was 22.5%, and reduced by almost half subsequently. However, the appearance of the Delta variant at the same time as the acceleration in the global vaccination campaign has created a new scenario, and at the moment it is not yet known whether it will cause a worsening or a relief for health systems^4,16,17^. The most remarkable conclusion of our study was that the patients admitted during the fifth wave were considerably younger than before, with an average age of 38.2 years, and mortality fell considerably to just 2.0%. Likewise, while patients in the early periods were admitted mostly to the Internal Medicine department, with average stays of more than 12 days, in the fifth they were treated mainly in the Emergency Department and the average stay reduced to 5.4 days. In addition, the percentage of patients moved to the Intensive Care Unit decreased significantly, from 6-10% to 2% and the percentage of patients requiring assisted respiratory intervention also decreased. The most common symptoms in all waves were fever, respiratory distress, and cough. Patients in the fifth wave had fewer comorbidities than those before, which is probably explained by the age difference. Our results are an empirical verification of the predictions previously drawn up in mathematical models that concluded that vaccination would markedly reduce the risk of Delta variant-associated COVID-19 resurgence and adverse outcomes. This is also backed up by data from South Korea^18^, the United States^9^, Germany^19^, and The Netherlands^20^. In contrast to that, the explosion of the Delta variant has been linked to a large increase in adverse events and deaths in India, where the vaccination campaign has been delayed by enormous logistical difficulties^21^.

We next investigated the factors associated with the death of patients, but results should be treated with caution since only 4 patients died, making any statistical analysis rather weak. That said, the qualitative characteristics of the deceased patients coincided with what is currently known about the risk factors for death from COVID-19^11^. The patients who died were much older than those who survived, and were affected more commonly by age-related comorbidities (Table 2). Moreover, the older patients were hospitalized for longer and more frequently needed assisted ventilation. It is noteworthy that these characteristics are more descriptive of patients in waves 1-3 (Table 3) than of wave 5.

**Table 2.** Differences between the clinical and demographic characteristics of the fifth wave patients in terms of whether they died or not

|  | Deceased |  |  |
| --- | --- | --- | --- |
| Variable | No<br>(n = 196) | Yes<br>(n = 4) | P-value |
| Demographic variables |  |  |  |
| Sex, female | 98 (50.0) | 2 (50.0) | 0.689 |
| Age | 37.7 (1.6) | 66.0 (10.6) | 0.012 |
| Smokers | 13 (6.6) | 0 (0.0) | 0.763 |
| Alcohol drinking habit | 15 (7.6) | 1 (25.0) | 0.285 |
| Symptoms |  |  |  |
| Fever | 108 (55.1) | 2 (50.0) | 0.610 |
| Respiratory distress | 52 (26.5) | 3 (75.0) | 0.064 |
| Pneumonia | 48 (24.5) | 2 (50.0) | 0.495 |
| Cough | 90 (45.9) | 1 (25.0) | 0.381 |
| Diarrhea | 19 (9.7) | 0 (0.0) | 0.669 |
| Chills | 5 (2.6) | 0 (0.0) | 0.903 |
| Acute kidney failure | 4 (2.0) | 0 (0.0) | 0.922 |
| Acute respiratory distress syndrome | 5 (2.6) | 1 (25.0) | 0.116 |
| Vomiting | 13 (6.6) | 0 (0.0) | 0.763 |
| Other symptoms* | 42 (21.4) | 0 (0.0) | 0.386 |
| Asymptomatic | 24 (12.2) | 1 (25.0) | 0.416 |
| Days of admission | 2.1 (4.7) | 18.7 (9.9) | < 0.001 |
| Comorbidities |  |  |  |
| Cardiovascular disease | 38 (19.4) | 3 (75.0) | 0.028 |
| Type 2 diabetes mellitus | 17 (8.7) | 2 (50.0) | 0.046 |
| Chronic neurological disease | 13 (6.6) | 1 (25.0) | 0.254 |
| Chronic lung disease | 13 (6.6) | 3 (75.0) | 0.002 |
| Chronic kidney disease | 7 (3.6) | 0 (0.0) | 0.866 |
| Cancer | 15 (7.6) | 1 (25.0) | 0.285 |
| Concomitant infectious disease | 17 (8.7) | 0 (0.0) | 0.699 |
| Chronic liver disease | 6 (3.1) | 0 (0.0) | 0.884 |
| Pregnancy | 4 (2.0) | 0 (0.0) | 0.922 |
| Postpartum | 2 (1.0) | 0 (0.0) | 0.960 |
| Respiratory intervention |  |  |  |
| Conventional oxygen therapy | 39 (19.9) | 3 (75.0) | 0.030 |
| Invasive mechanical ventilation | 5 (2.6) | 1 (25.0) | 0.116 |
| High-flow oxygen therapy | 11 (5.6) | 0 (0.0) | 0.796 |
| Non-invasive mechanical ventilation | 2 (1.0) | 1 (25.0) | 0.059 |

\* Anosmia, ageusia, odynophagia, headache, anorexia, hyporexia, myalgia, and arthralgia. Results are given as numbers and percentages or as means and standard errors.

**Table 3.**
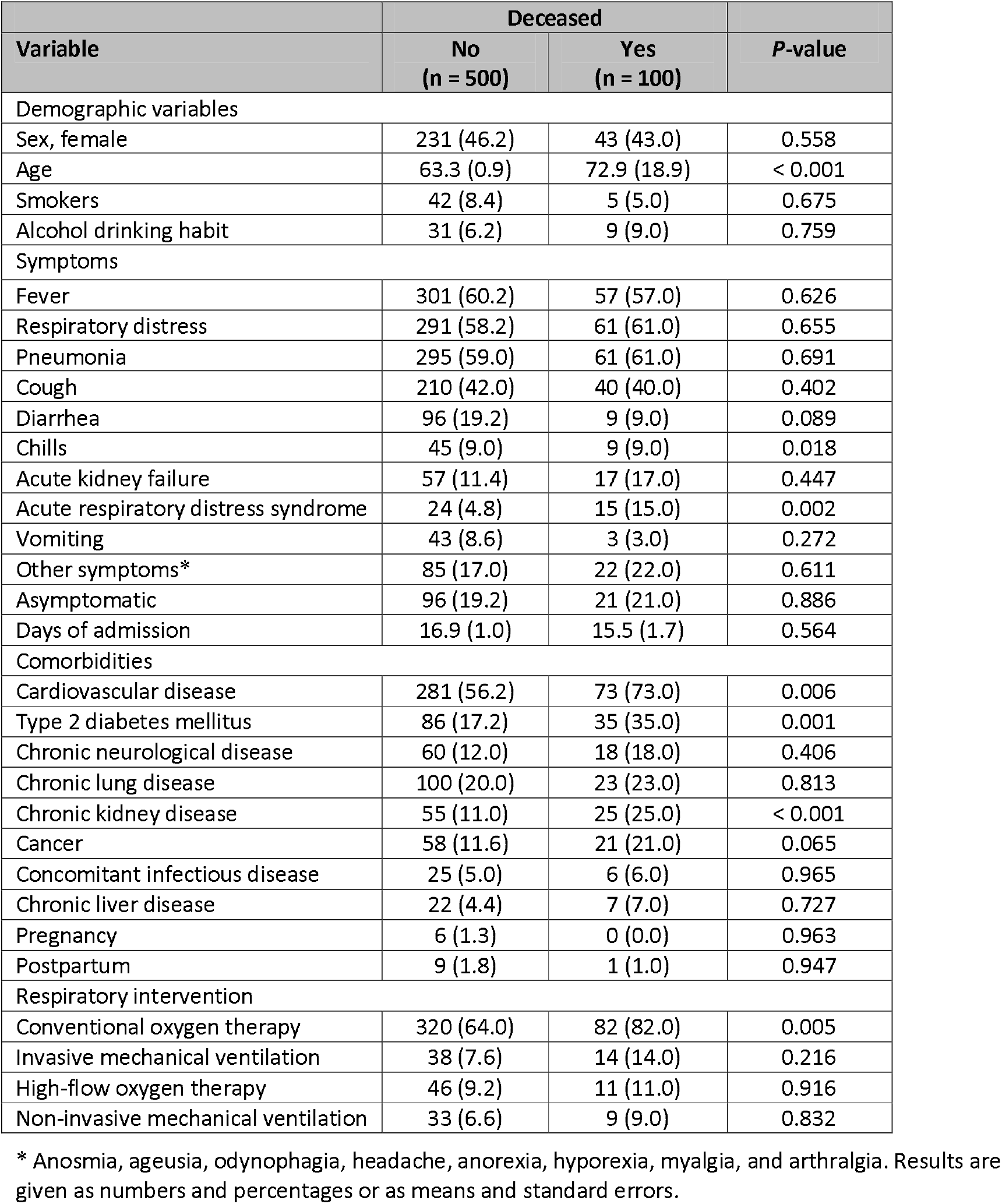
Differences between the clinical and demographic characteristics of the patients belonging to the waves 1-3 in terms of whether they died or not

| Variable | Deceased |  | P-value |
| --- | --- | --- | --- |
|  | No<br>(n = 500) | Yes<br>(n = 100) |  |
| Demographic variables |  |  |  |
| Sex, female | 231 (46.2) | 43 (43.0) | 0.558 |
| Age | 63.3 (0.9) | 72.9 (18.9) | < 0.001 |
| Smokers | 42 (8.4) | 5 (5.0) | 0.675 |
| Alcohol drinking habit | 31 (6.2) | 9 (9.0) | 0.759 |
| Symptoms |  |  |  |
| Fever | 301 (60.2) | 57 (57.0) | 0.626 |
| Respiratory distress | 291 (58.2) | 61 (61.0) | 0.655 |
| Pneumonia | 295 (59.0) | 61 (61.0) | 0.691 |
| Cough | 210 (42.0) | 40 (40.0) | 0.402 |
| Diarrhea | 96 (19.2) | 9 (9.0) | 0.089 |
| Chills | 45 (9.0) | 9 (9.0) | 0.018 |
| Acute kidney failure | 57 (11.4) | 17 (17.0) | 0.447 |
| Acute respiratory distress syndrome | 24 (4.8) | 15 (15.0) | 0.002 |
| Vomiting | 43 (8.6) | 3 (3.0) | 0.272 |
| Other symptoms* | 85 (17.0) | 22 (22.0) | 0.611 |
| Asymptomatic | 96 (19.2) | 21 (21.0) | 0.886 |
| Days of admission | 16.9 (1.0) | 15.5 (1.7) | 0.564 |
| Comorbidities |  |  |  |
| Cardiovascular disease | 281 (56.2) | 73 (73.0) | 0.006 |
| Type 2 diabetes mellitus | 86 (17.2) | 35 (35.0) | 0.001 |
| Chronic neurological disease | 60 (12.0) | 18 (18.0) | 0.406 |
| Chronic lung disease | 100 (20.0) | 23 (23.0) | 0.813 |
| Chronic kidney disease | 55 (11.0) | 25 (25.0) | < 0.001 |
| Cancer | 58 (11.6) | 21 (21.0) | 0.065 |
| Concomitant infectious disease | 25 (5.0) | 6 (6.0) | 0.965 |
| Chronic liver disease | 22 (4.4) | 7 (7.0) | 0.727 |
| Pregnancy | 6 (1.3) | 0 (0.0) | 0.963 |
| Postpartum | 9 (1.8) | 1 (1.0) | 0.947 |
| Respiratory intervention |  |  |  |
| Conventional oxygen therapy | 320 (64.0) | 82 (82.0) | 0.005 |
| Invasive mechanical ventilation | 38 (7.6) | 14 (14.0) | 0.216 |
| High-flow oxygen therapy | 46 (9.2) | 11 (11.0) | 0.916 |
| Non-invasive mechanical ventilation | 33 (6.6) | 9 (9.0) | 0.832 |

\* Anosmia, ageusia, odynophagia, headache, anorexia, hyporexia, myalgia, and arthralgia. Results are given as numbers and percentages or as means and standard errors.

It is well known that the efficacy of vaccines depends largely on whether the full or only partial regimen has been administered, or indeed the time elapsed from vaccination to infection with SARS-CoV-2. For example, a study in Canada has reported that vaccine effectiveness against Delta after partial vaccination was lower compared to Alpha with both the Moderna (72% vs. 83%) and the Pfizer (56% vs. 66%) vaccines, while full vaccination increased protection to comparable levels (87%)^22^. In addition, a British study reported that effectiveness was lower against Delta after one dose of Pfizer or Astra-Zeneca vaccines compared to Alpha (30.7% vs. 48.7%), and increased after the second dose, although it was always lower for the Delta variant^23^. Considering that, we wanted to study the vaccination status of the patients who died in the fifth wave of our study. Most of the patients admitted during the fifth wave were not vaccinated. One hundred and forty-two surviving patients (71.9%) had not received any dose of vaccine and 55 (28.1%) had received at least one dose. We were surprised to find that 3 of the 4 patients who died had been partially or fully vaccinated. One of them was a male over 70 years of age who had received both doses of the Pfizer vaccine between March and April 2021, and died on early August. The time elapsed to build immunity had been sufficient but the patient was older and he also suffered from epilepsy. The most plausible explanation for his death is that he did not have a sufficiently effective immune response. Another victim was a woman in her thirties who had received a single dose of Astra-Zeneca vaccine at the end of June and died from bilateral pneumonia in mid July, not having had time to receive a second dose. Another patient who died was a man over 60 years old who had received a single dose of the Pfizer vaccine at the beginning of May and died at the end of July. This patient suffered a chronic obstructive pulmonary disease prior to infection and died of bilateral pneumonia. It seems therefore that the unfortunate outcome of two of these patients might be explained by their advanced age and associated comorbidities.

In conclusion, our results show a marked decrease in the severity of infected patients, in the number of admissions to the Intensive Care Unit, and in the number of deaths, too, during the fifth wave of the pandemic in Spain. This was linked to the relative youth of the patients and the absence of comorbidites. This improvement coincided with the majority of the population, and especially all the elderly, having been vaccinated, despite the explosion of the Delta variant. A caveat to the present study is its single-center nature in a specific region of Mediterranean Europe. However, we think that the results can be extrapolated to regions and countries that have had similar circumstances, i.e. a high incidence of the Delta variant together with a high vaccination rate. At a time when the elderly population is already largely protected, a special effort must be made to vaccinate young people in order to achieve group immunity as soon as possible.

## Data Availability

All data produced in the present work are contained in the manuscript and its supplementary files.

## Data availability

Supporting datasets are available as Supplementary Information (S1_File.xls).

## Acknowledgments

Editorial assistance and English language correction was provided by Phil Hoddy at the Service of Linguistic Resources of the *Universitat Rovira i Virgili*.

## Author contributions

S.I.: Study supervision. Concept and study design. Data collection. Data interpretation. Drafting, revision and final approval of manuscript. A.F.L.A.: Concept and study design. Data interpretation. Revision and final approval of manuscript. M.J.L.O.: Data collection. Revision and final approval of manuscript. A.H.A.: Data collection. Revision and final approval of manuscript. S.S.M.: Data collection. Revision and final approval of manuscript. J.J..: Data collection. Revision and final approval of manuscript. J.C.: Study supervision. Concept and study design. Data analysis and statistics. Data interpretation. Drafting, revision and final approval of manuscript. A.C.: Study supervision. Concept and study design. Data interpretation. Revision and final approval of manuscript.

